# Quantifying Reproducibility of Common Laboratory Tests in a Tertiary Care Hospital for More Realistic Synthetic Data in Machine Learning

**DOI:** 10.64898/2026.10.20.26366057

**Authors:** Dawson Dean

## Abstract

Data from medical records are often limited in size, creating challenges for Artificial Intelligence training, and an opportunity for data augmentation techniques to create synthetic data. A natural question is how much data can be randomized and still be realistic. The main goal of this study is to attempt to measure the reproducibility of labs drawn in practice, so these ranges can be used for generating more realistic synthetic data. A second goal is to inform clinicians of the real error ranges of their laboratory data so these values may be better interpreted.

## 1. Introduction

Data from medical records are often limited in size, creating challenges for Artificial Intelligence training. As a result, training sets are often expanded with synthetic data created by data augmentation techniques like SMOTE [1], Generative Adversarial Networks, Noise Injection and more[2]. A natural question is how much data can be randomized and still be realistic and useful for training. Simply randomizing within a range, such as 5 or 10 percent, may not reflect the behavior of real labs. A more medically realistic answer is to randomize within the limits of normal laboratory precision. Besides the actual laboratory analysis, the blood collection process can also introduce significant error. As a result, the precision specifications of commercial lab testing equipment is not enough, we need empiric analysis. For example, a peripheral blood test may be drawn from a fresh venous puncture or else drawn through an existing peripheral IV which can cause hemolysis. Once collected, the blood may be diluted by saline flush or exposed to various temperature differences and time delays. The primary goal of this study is to measure the accuracy of labs drawn in practice, so these ranges can be used for generating synthetic data to augment training. A second goal is to inform clinicians of the real error ranges of their laboratory data so these values may be better interpreted.

### 2. Materials

This study is a retrospective analysis of 194,905 patients across 18,097,846 admissions and clinic visits at the University of Kentucky Hospitals, which are teaching hospitals serving central and eastern Kentucky in the USA. All patients were admitted to general Internal Medicine Teams between 2020 and 2025 inclusive, and all patients were at least 18 years old. All data analysis was performed with Python software running on Python V3.7[3] using standard numerical libraries. The source code for all analysis is open source and may be downloaded at http://www.dawsondean.com/publicSrcCode/LabReproducibility_SrcCode.tar

This study identified pairs of the same lab value collected within fifteen minutes apart and compares their difference. Comparing the results from these near simultaneous tests suggests the reproducibility of the overall test and may be indicative of its accuracy. Fifteen minutes is too short of a time for the first lab result to be available in the electronic medical record, so the second collection is often inadvertent redundancy rather than a response to an obviously wrong lab. Such redundancy may occur because some labs appear in separate tests. For example, sodium or creatinine are part of a Basic Metabolic Panel, a Comprehensive Metabolic panel, and a Renal Function Panel, and a clinician may order two of these same panels and get the same lab twice. Alternatively, duplicate lab tests may be ordered if a patient is moving between two different services, such as the Emergency Department and an Internal Medicine ward unit. This study does not equate two labs that may be drawn from different types of serum. For example, a sodium value from arterial blood that is part of an Arterial Blood Gas panel, and a sodium value from venous blood that is part of a Comprehensive Metabolic Panel are considered different labs, even if they are collected minutes apart.

This study involves human participants but University of Kentucky IRB exempted this study. All patients were de-identified, and an application to Institution Review Board (IRB) was submitted and determined exempt because the study only analyzed de- identified anonymous data.

## 3. Results

Table 1 describes the number of pairs of labs collected within 15 minutes of each other, and the maximum difference between 90% of these pairs, maximum difference between 95% of these pairs

**Table 1.** Pairs of Lab Identical Values Collected Within 15 Minutes.

| Lab | Number of Pairs | Max Difference for 90% of Pairs | Max Difference for 95% of Pairs | Normal Range | Units |
| --- | --- | --- | --- | --- | --- |
| Hemoglobin (Hgb) | 1419 | 0.7 | 1.2 | 11.2 - 15.7 | g/dL |
| Platelets | 914 | 28 | 45 | 155 - 369 | 1000/uL |
| White Blood Cells | 1126 | 0.95 | 1.55 | 3.7 - 10.3 | 1000/uL |
| Sodium | 1519 | 3.0 | 4.0 | 136 - 145 | mmol/L |
| Potassium | 1448 | 0.5 | 0.8 | 3.6 - 4.9 | mmol/L |
| Bicarbonate | 1208 | 1.55 | 1.55 | 22 - 29 | mmol/L |
| Creatinine | 1388 | 0.11 | 0.12 | 0.6 - 1.1 | mg/dL |
| Calcium | 1223 | 0.4 | 0.6 | 8.9 - 10.2 | mg/dL |
| Phosphorus | 490 | 0.7 | 1.0 | 2.5 - 4.5 | mg/dL |
| Magnesium | 469 | 0.2 | 0.4 | 1.9 - 2.4 | mg/dL |
| Alanine<br>Aminotransferase<br>(ALT) | 370 | 4 | 9 | 10 - 35 | U/L |
| Aspartate<br>Aminotransferase<br>(AST) | 372 | 8 | 15 | 10 - 35 | U/L |
| Alkaline<br>Phosphatase<br>(ALP) | 381 | 8 | 15 | 35 - 104 | U/L |
| Total Bilirubin | 356 | 0.1 | 0.3 | 0.2 - 1.1 | mg/dL |
| Total Protein | 519 | 0.5 | 0.6 | 6.3 - 7.9 | g/dL |
| Albumin | 713 | 0.2 | 0.3 | 3.5 - 5.2 | g/dL |
| C-Reactive<br>Protein | 31 | 1.55 | 1.55 | Below 8.0 | mg/dL |

The value distributions for Hemoglobin, Potassium and Creatinine are shown below.

**Figure 1.**
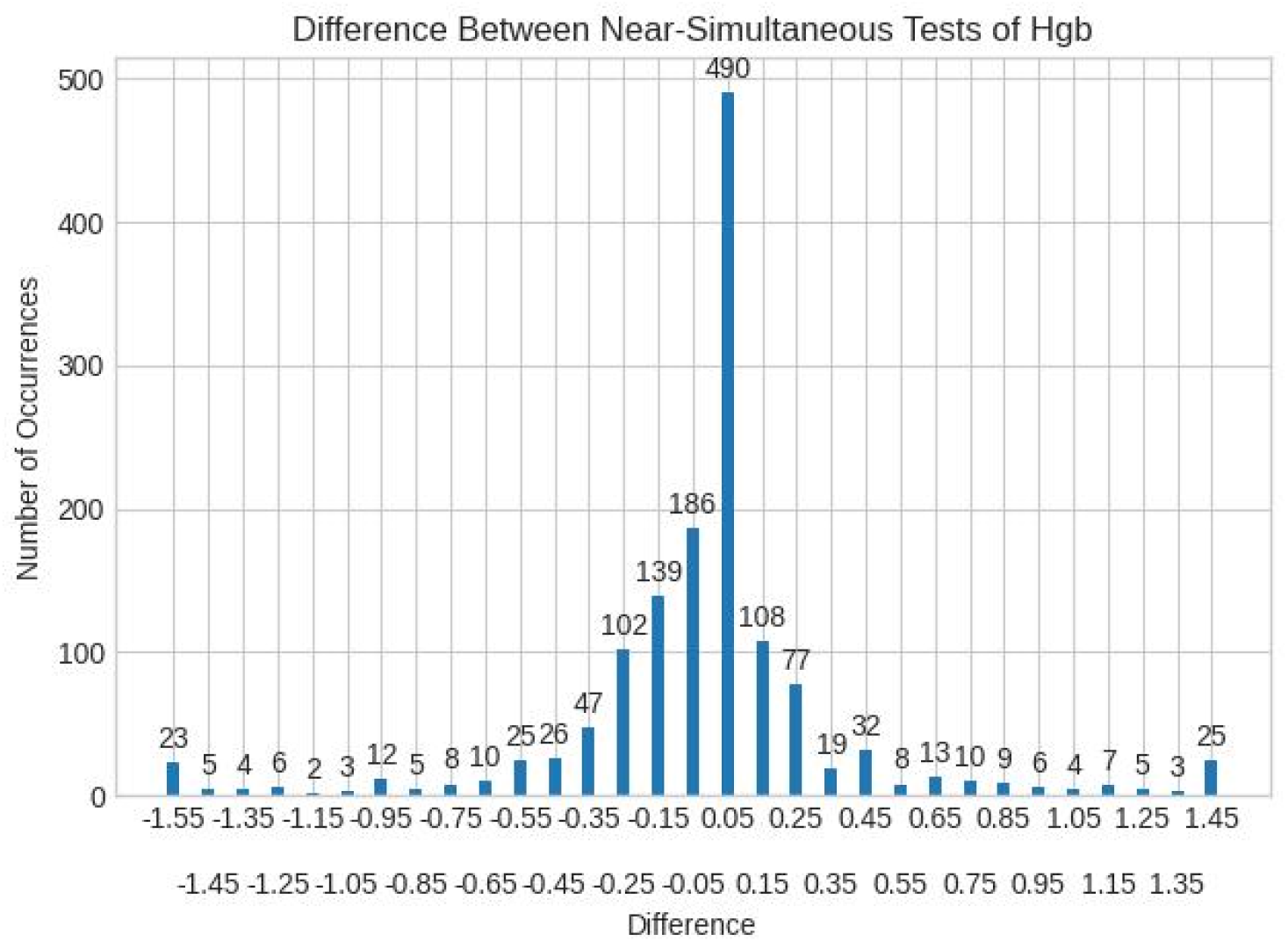
Distribution of Differences Between Hemoglobin Values Collected Within Fifteen Minutes

**Figure 2.**
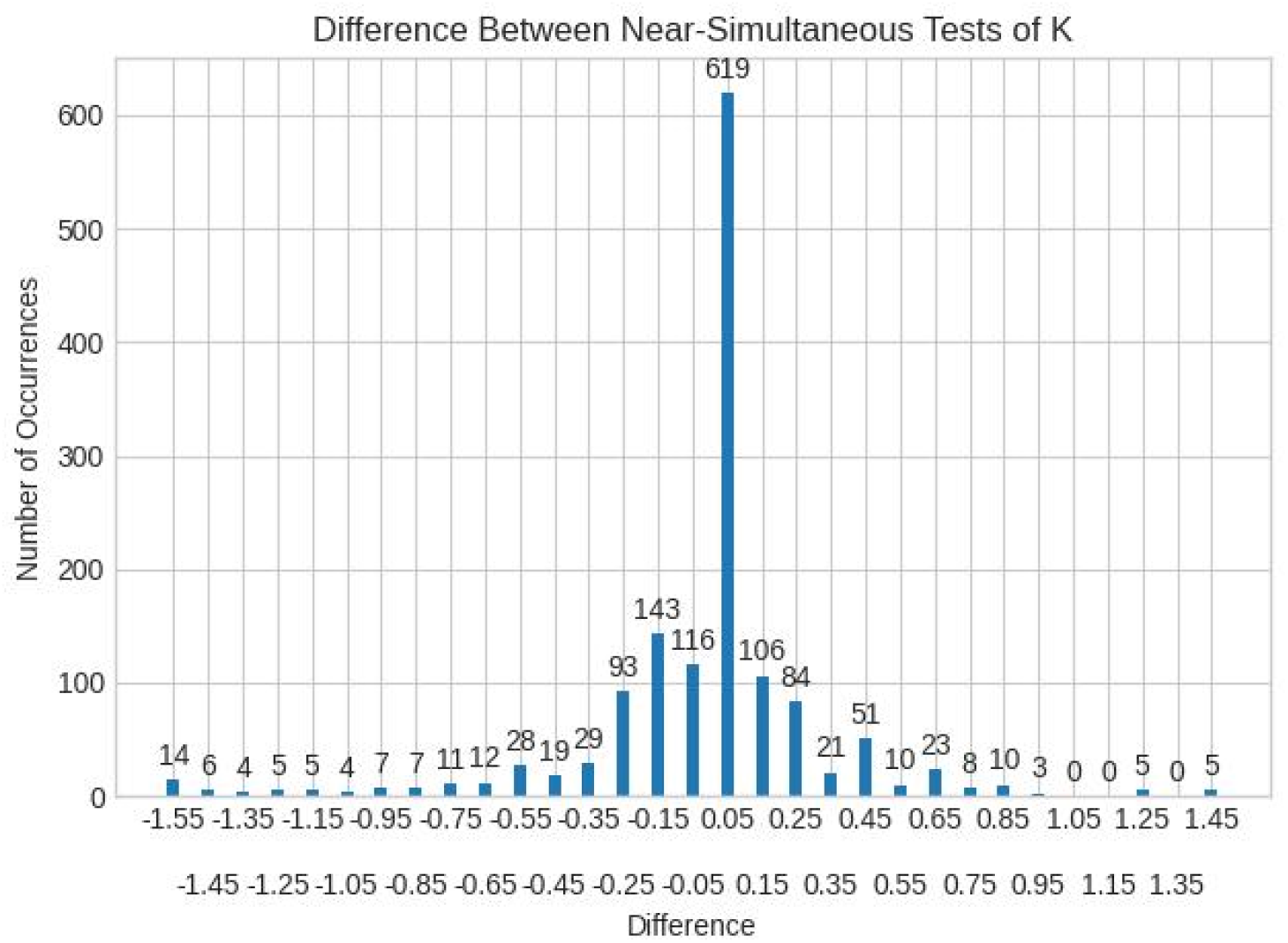
Distribution of Differences Between Potassium Values Collected Within Fifteen Minutes

**Figure 3.**
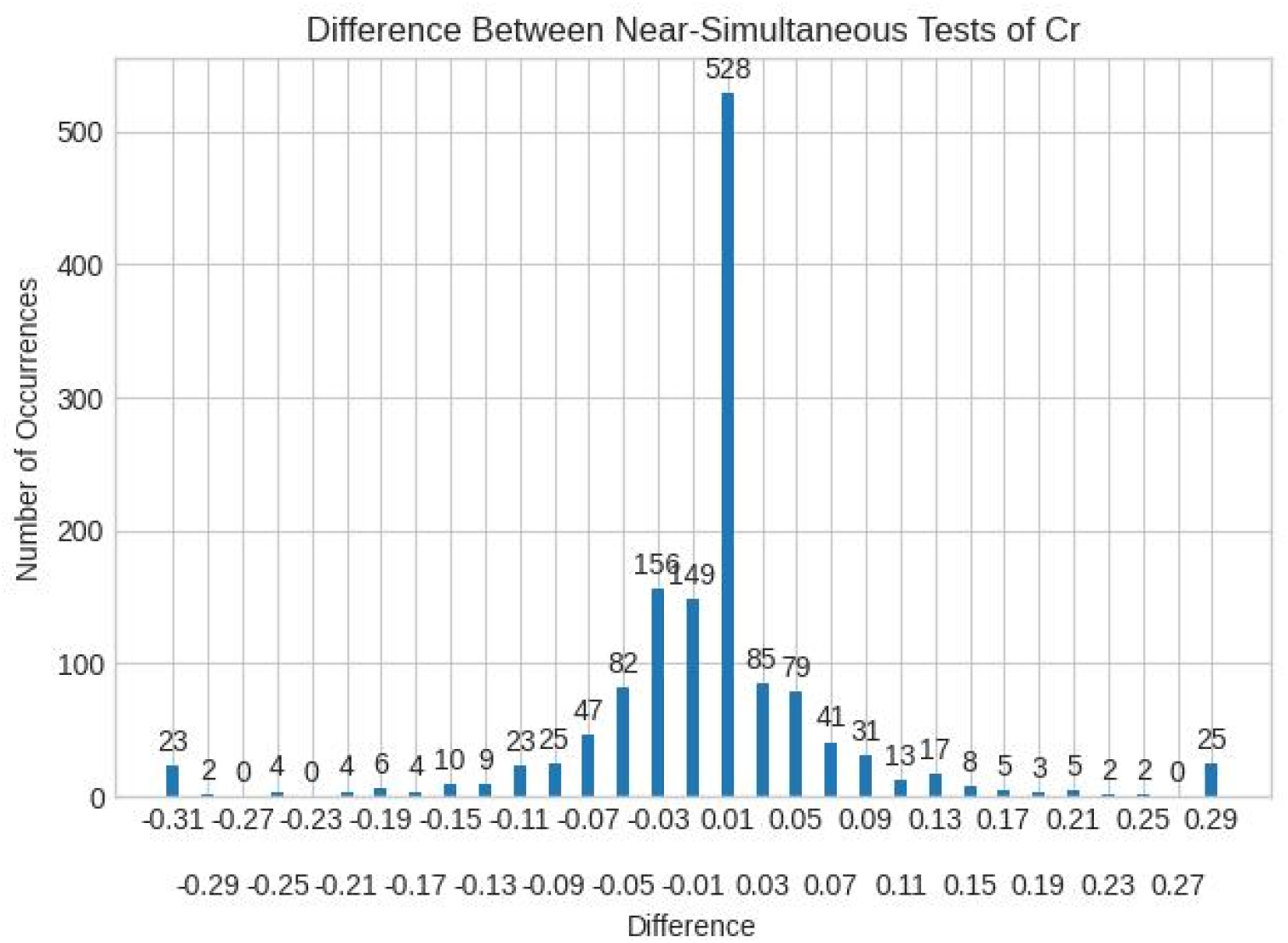
Distribution of Differences Between Creatinine Values Collected Within Fifteen Minutes

## 4. Discussion

Most of these blood tests demonstrate good reproducibility, for example 88% of Hemoglobin pairs were within 0.5 g/dL, and no sodium pairs were more than 7 mmol/L apart. However, the relative difference depends on the scale of a normal value. For example, a difference of 0.8 mmol/L for potassium represents 20% of a normal value of 4.0 mmol/L, while a difference of 4 mmol/L for sodium values represents 2% of a normal sodium of 140 mmol/L. Additionally, accuracy varies with the lab, and some contaminant processes may affect some labs (such as serum potassium) more than others. For example, hemolysis during blood collection ruptures red blood cells and releases ions that are normally much more highly concentrated intracellularly. This means that simple noise injection techniques should treat different variables differently; and adjust the range of artificial noise according to the empiric normal variance in each lab.

Beyond software development, these differences can also have clinical significance at the bedside. For example, 95% of Hemoglobin pairs are within 1.2 g/dL of each other, but that means that a patient’s whose blood Hemoglobin drops 1.0 g/dL overnight may still be within expected error. For Sodium, 95% of Hemoglobin pairs are within 4.0 mmol/L of each other, but guidelines for correcting Hyponatremia [4] recommend increasing serum sodium no more than 8 mmol/L in a day. The error margin of 4 mmol/L is 50% of this desired total daily increase.

This paper has several strengths: it uses empiric observations to measure reproducibility, and therefore to infer accuracy. There are however several weaknesses: this study did not account for possible confounding factors, such as other electrolyte concentrations in serum and the possible interference with measurements, such as pseudohyponatremia. The study also did not consider other medical conditions, such as intravascular hemolysis processes like Disseminated Intravascular Coagulation or Tumor Lysis Syndrome.

The project described was supported by the NIH National Center for Advancing Translational Sciences through grant number UL1TR001998. The content is solely the responsibility of the authors and does not necessarily represent the official views of the NIH.

## Data Availability

Data are available upon reasonable request. The data that support the findings of this study are available on request from the corresponding author, [DD] pending IRB approval. The data are not publicly available
due to their containing information that could compromise the privacy of research participants

